# Efficacy of remdesivir in COVID-19 patients with a simulated two-arm controlled study

**DOI:** 10.1101/2020.05.02.20088559

**Authors:** Chen-Yang Hsu, Chao-Chih Lai, Amy Ming-Fang Yen, Sam Li-Sheng Chen, Hsiu-Hsi Chen

## Abstract

While the recent study on the compassionate use of remdesivir for COVID-19 patients has shown a 68% clinical improvement^7^ it is a one-arm study that renders the evaluation of the efficacy in reducing death and the length of stay of hospitalization intractable due to a lacking of the control group. We came up with a two-arm controlled study design to simulate the treated and the untreated (control group) group by applying two respective transition models to the empirical data on dynamics of the disease severity (Figure 2 of the original article^7^) that are classified into low- (no and low oxygen supplement), medium- (non-invasive ventilator and high oxygen supplement), and high-(ECMO and invasive ventilator) from enrolment until discharge, death or the end of follow-up. By using a simulated two-arm controlled study, the remdesivir treatment group as opposed to the control group led to a statistically significantly 29% (95% CI: 22-35%) reduction of death from COVID-19. The treated group also revealed a 33% (95% CI 28-38%) significantly higher odds of discharge than the control group. The median time to discharge for the treated group (5.5 days, 16.5 days, and 29.5 days for low-, medium-, and high-risk state, respectively) was around half of those of the control arm. Our results with a simulated two-arm controlled study have not only corroborated the efficacy of remdesivir but also made great contribution to designing a further large-scale randomized controlled trial. They have significant implications for reducing transmission probability and infectious time of COVID-19 patients when contacting with susceptible health care workers during hospitalization.

**Key Points:** *Question:* What is the efficacy of remdesivir in reducing advanced disease state or death from COVID-19 and the length of stay of hospitalization?

*Findings:* Remdesivir treatment results in a 33% significantly higher odds of discharge, a 29% significantly lower risk of death, and a 39% significantly lower risk for the combined endpoint of severe status and death. The median time to discharge for the remdesivir treated group was around half of the median time-to-discharge compared with the control arm.

*Meaning:* Remdesivir is effective in treating COVID-19 patients in terms of enhancing recovery and accelerating discharge.

## Introduction

As of April 25^th^, 199,272 deaths out of 2,840,830 confirmed COVID-19 cases, amounting to 7% case-fatality, have been noted during COVID-19 pandemic. Besides, the recovery rate is also extremely low to less than 30%.^1^ To reduce its death and the length of stay (LOS) in hospitalization resulting from COVID-19, the administration of anti-viral therapy may provide a solution to treating these emerging infected cases.^2–6^ The recent study on the compassionate use of remdesivir for patients with COVID-19,^7^ albeit it is a one-arm before-and-after design, has provided a golden opportunity to offer a simulated two-arm controlled study for evaluation of the efficacy in reducing advanced disease state or death from COVID-19 and the length of stay of hospitalization.

## Material and Methods

The empirical data used for building up the simulated two-arm controlled study and the following analysis on the efficacy of remdesivir were derived from a recent study on the compassionate use of remdesivir for 53 patients with COVID-19 as shown in Figure 2 of the original article that gives a clear profile of dynamics of disease transition according to the severity of disease from the date of being administered by remdesivir until discharge, death, or the end of study.^7^ The risk state of these patients can be classified into low- (no and low oxygen supplement), medium- (non-invasive ventilator and high oxygen supplement), and high-(ECMO and invasive ventilator) risk states. The aggregated data on each transition mode are listed in **Table 1** after the translation from the data provided in the original published article.^7^

**Table 1.** Empirical data on the evolution of 53 COVID-19 patients translated from published data.^7^

| Disease Evolution |  | Frequency | (%) |
| --- | --- | --- | --- |
| From | To |  |  |
| On Enrolment |  |  |  |
|  | Low | 12 | (22.6) |
|  | Medium | 7 | (13.2) |
|  | High | 34 | (64.2) |
| During Follow-up |  |  |  |
| Low | Low | 197 | (87.9) |
|  | Medium | 4 | (1.8) |
|  | Discharge | 23 | (10.3) |
| Medium | Low | 12 | (9.1) |
|  | Medium | 113 | (85.6) |
|  | High | 5 | (3.8) |
|  | Discharge | 1 | (0.8) |
|  | Death | 1 | (0.8) |
| High | Low | 11 | (2.5) |
|  | Medium | 11 | (2.5) |
|  | High | 403 | (93.3) |
|  | Discharge | 1 | (0.2) |
|  | Death | 6 | (1.4) |

**Figure 1 (A) and (B)** shows two transition models between the risk state of disease and also the final destinations of both discharge and death. **Figure 1 (B)** models the forward progression, leading to recovery and death, without the use of remdesivir whereas **Figure 1 (A)** models not only forward progression but also backward regression resulting from the use of remdesivir.

**Figure 1.**
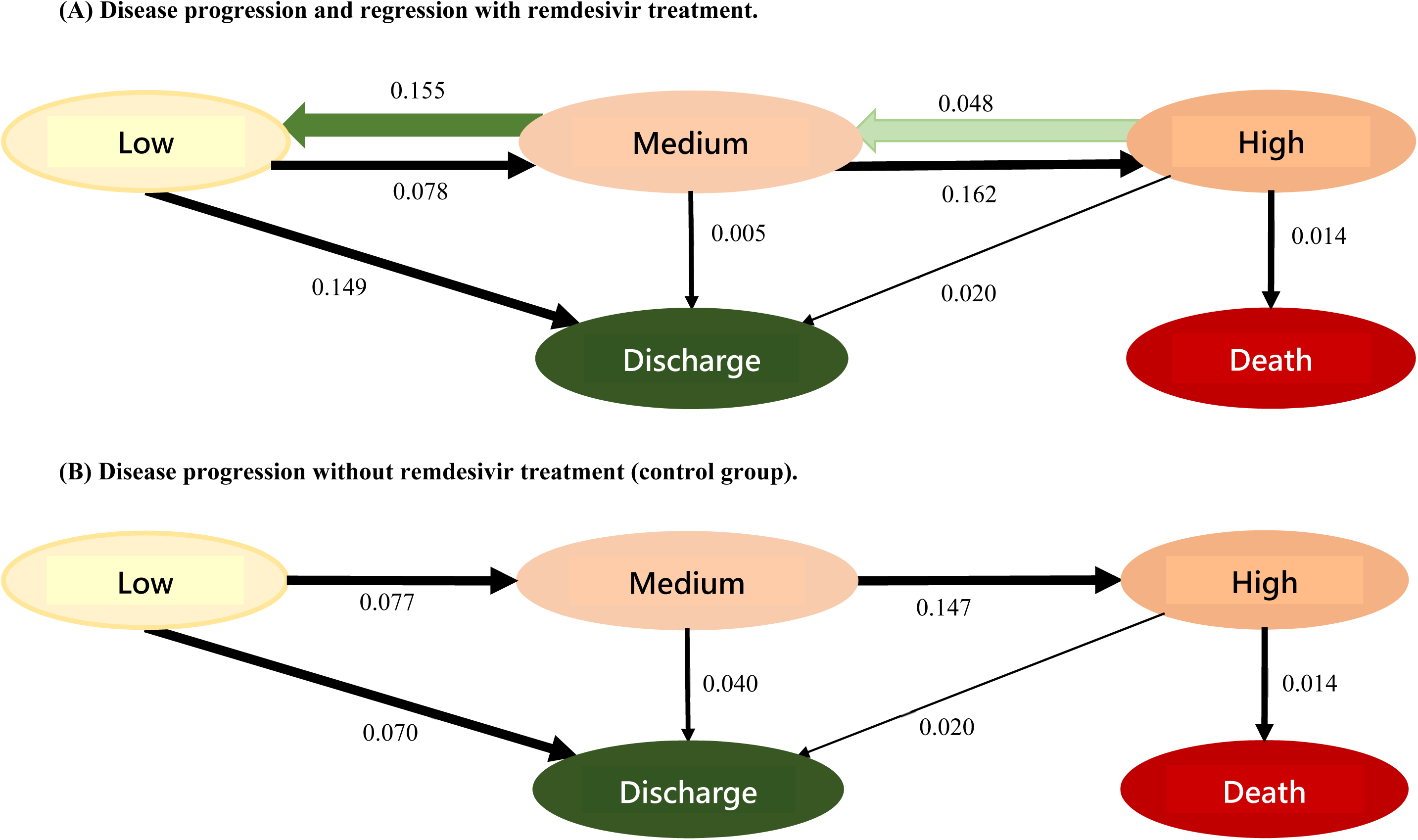
Two disease transition models for COVID-19 patients.

The estimates derived from the model without the use of remdesivir **(Figure 1 (B))** were applied to simulating a pseudo-control group. The treated group with remdesivir that was also simulated in the light of the corresponding estimates of **Figure 1 (A)**. This simulated treated group had an adequate fit with the observed data with the administration of remdesivir (P=0.38). Such a two-arm controlled study, which is equivalent to a randomized controlled trial, was therefore simulated and used for evaluating the efficacy of remdesivir treatment in accelerating discharge and decelerating subsequent deaths.

## Results

The daily rate of transition between risk states, discharge, and death were estimated and analyzed by applying a continuous-time Markov model^8^ with Bayesian Markov Chain Monte Carlo (MCMC) simulation in the light of likelihood functions based on aggregated data as shown in **Table 1**. As shown in **Figures 1(A)** and **(B)** with the corresponding daily transition rates, the efficacy of remdesivir in reducing subsequent progression to high-risk state and death was mainly attributed to the two daily regression rates estimated as 0.048 (95% credible interval (CI): 0.027-0.066) from medium-risk to low-risk state and estimated as 0.155 (95% CI: 0.098-0.227) from high-risk to medium-risk state during hospitalization. The higher overall discharge rate regardless of risk states in the treated group as opposed to the control group is mainly explained by the mechanism of two regression rates during hospitalization, resulting in a double proportion of discharge in the low-risk state in the treated group as opposed to the control group (0.149 (95% CI: 0.081-0.206) vs 0.070 (95% CI: 0.030-0.111)). This also accounts for why the rate of discharge in the treated group was lower for the medium-risk state than the control group because a substantial proportion of patients regressing from medium-risk to low-risk state in the treated group during hospitalization. The similar logic is applied to the identical discharge rate between the two groups given the modest regression rate from high-risk to medium-risk state.

**Figure 2** shows the cumulative probabilities of discharge **(Figure 2 (A))** and death **(Figure 2(B))** by the two treatment groups during the 28-day follow-up since enrolment. A higher probability of discharge and a lower probability of death were noted for the treated group (green). The use of remdesivir led to a statistically significantly 29% reduction of death (relative risk: 0.71, 95% CI: 0.65-0.78). When the high-risk state and death was combined as the endpoint, the corresponding reduction was still statistically significant up to 39% (relative risk: 0.61 (95% CI: 0.58-0.63)). The cumulative discharge probability was 58.6% (95% CI: 57.9-59.5%) for the remdesivir group and 44.1% (95% CI: 42.5-45.7%) for the control group, respectively, giving a 33% (95% CI 28-38%) significantly higher odds of discharge for the treated group.

**Figure 2.**
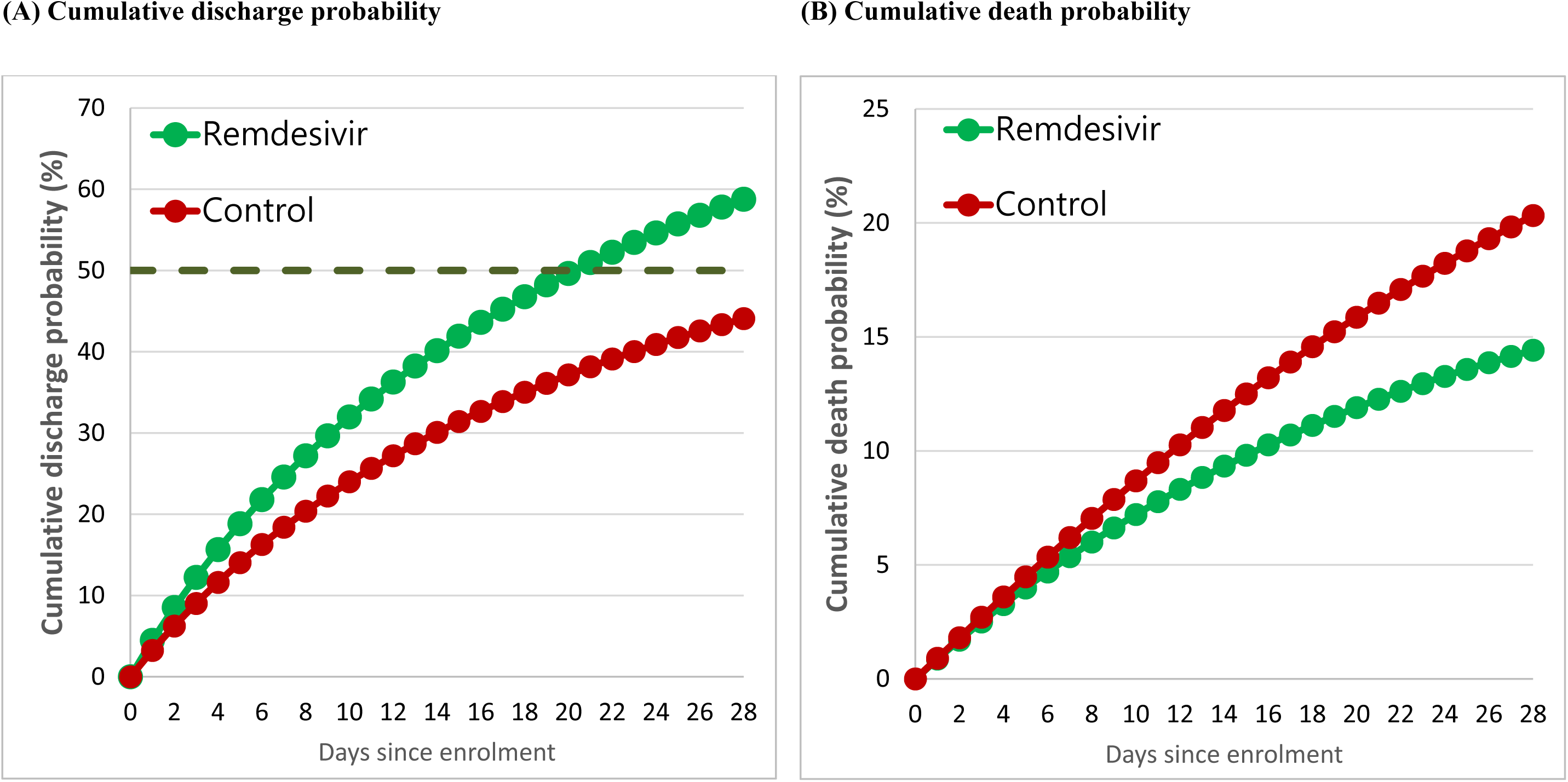
Cumulative rate of discharge and death by treatments.

The median time to discharge for the treated group was estimated as 5.5 days, 16.5 days, and 29.5 days for patients with low-, medium-, and high-risk state, respectively, which was around half of the median time-to-discharge compared with the control arm when the follow-up time was extended to two months.

## Discussion

The advantages of our secondary analysis of the empirical data from the original one-arm compassionate study on the use of remdesivir,^7^ while using the simulated two-arm controlled study, are three-fold. Firstly, we improved the weakness of a lacking control group in the original study as a proportion of COVID-19 patients may be discharged with the recovery dispensing with the use of remdesivir. Second, such a simulated two-arm study design enables us to evaluate the efficacy of remdesivir with clear primary endpoints including death and discharge rather than only based on the clinical improvement before and after the use of remdesivir defined by live discharge from hospital, a decrease of at least 2 points from baseline on the supplementary oxygen use, or both as used in the original article.^7^ Third, the results from such a simulated two-arm controlled study are supposed to be closer to those using a real two-arm randomized controlled trial in the near future in terms of intention-to-treat analysis principle. Such a simulated two-arm controlled study may also account for why the efficacy of reducing death and accelerating discharge in the current two-arm controlled study was approximately one-third whereas the efficacy of clinical improvement reported in the original one-arm study without the control group and intention-to-treat principle was two-thirds.

The results on the efficacy of remdesivir in reducing death and length of stay resulting from COVID-19 have also two significant implications for containing COVID-19 pandemic. Firstly, it reduces the sequelae of COVID-19 and also accelerates its recovery. Besides, the administration of remdesivir may also reduce transmission probability and also infectious time of the infective in contact with the susceptible subjects. Such an efficacy of prophylactic and therapeutic anti-viral therapy has been demonstrated in influenza.^9–10^

In conclusion, a simulated two-arm controlled study based on data from one-arm compassionate use of remdesivir demonstrates one-third reduction of death and half of median time-to-discharge attributable to COVID-19. The results would make contribution to designing a large-scale randomized controlled trial study such as sample size determination and also have significant implications for reducing transmission probability and infectious time of COVID-19 patients when contacting with susceptible health care workers during hospitalization.

## Data Availability

All data and codes are available from the author on request.

## Acknowledgements

This work was financially supported by the “Innovation and Policy Center for Population Health and Sustainable Environment (Population Health Research Center, PHRC), College of Public Health, National Taiwan University” from The Featured Areas Research Center Program within the framework of the Higher Education Sprout Project by the Ministry of Education (MOE) in Taiwan.

## Author contributions

Conceptualization: H.H.C. and C.C.L; study design: C.Y.H. and H.H.C.; methodology: A.M.F.Y., S.L.S.C., and H.H.C.; data retrieval and management: C.Y.H. and C.C.L.; statistical analysis: C.Y.H. and A.M.F.Y.; computer programming: C.Y.H., A.M.F.Y., and S.L.S.C.; writing: C.Y.H. and H.H.C.; revision of draft: H.H.C.

## Competing interests

Authors declare no competing interests

## Funding Support

Ministry of Science and Technology, Taiwan (MOST 107-3017-F-002-003; MOST 108-2118-M-002-002-MY3; MOST 108-2118-M-038-001-MY3; MOST 108-2118-M-038-002-MY3), Ministry of Education, Taiwan (NTU-107L9003).

## Role of Funder/Sponsor

The funders had no role in the design and conduct of the study; collection, management, analysis, and interpretation of the data; preparation, review, or approval of the manuscript; and decision to submit the manuscript for publication.

## Notes

### Competing Interest Statement

The authors have declared no competing interest.

### Funding Statement

All data and codes are available from author on request.

